# Sugar- or artificially-sweetened beverage consumption, physical activity, and risk of cardiovascular disease in US adults

**DOI:** 10.1101/2023.04.17.23288711

**Authors:** Lorena S. Pacheco, Deirdre K. Tobias, Yanping Li, Shilpa N. Bhupathiraju, Walter C. Willett, David S. Ludwig, Cara B. Ebbeling, Danielle E. Haslam, Jean-Philippe Drouin-Chartier, Frank B. Hu, Marta Guasch-Ferré

**Affiliations:** Department of Nutrition, Harvard T.H. Chan School of Public Health, Boston, MA, USA; Division of Preventive Medicine, Department of Medicine, Brigham and Women’s Hospital and Harvard Medical School, Boston, MA, USA; Channing Division of Network Medicine, Department of Medicine, Brigham and Women’s Hospital, Harvard Medical School, Boston, MA, USA; Department of Epidemiology, Harvard School of Public Health, Boston, MA, USA; New Balance Foundation Obesity Prevention Center, Boston Children’s Hospital, Boston, MA, USA; Faculté de Pharmacie, Université Laval, Quebec City, Quebec, Canada; Centre Nutrition Santé et Societé (NUTRISS), Institut Sur la Nutrition et les Aliments Fonctionnnels (INAF), Université Laval, Quebec City, Quebec, Canada; Department of Public Health and Novo Nordisk Foundation Center for Basic Metabolic Research, Faculty of Health and Medical Sciences, University of Copenhagen, Copenhagen, Denmark

**Keywords:** joint association, sucrose, aspartame, sugary beverages, diet drinks, coronary heart disease, stroke

## Abstract

**Background:** The extent to which physical activity attenuates the detrimental effects of sugar (SSBs)- or artificially-sweetened beverages (ASBs) on the risk of cardiovascular disease is unknown.

**Methods:** We used Cox proportional-hazards models to calculate hazard ratios and 95% confidence interval [HR (CI)] between SSB or ASB intake and physical activity with cardiovascular disease risk among 65,730 women in the Nurses’ Health Study (1980-2016) and 39,418 men in the Health Professional’s Follow-up Study (1986-2016), who were free from chronic diseases at baseline. SSBs and ASBs were assessed every 4-years and physical activity biannually.

**Results:** A total of 13,269 cardiovascular events were ascertained during 3,001,213 person-years of follow-up. Compared with those that never/rarely consumed SSBs or ASBs, HR and 95% CI for cardiovascular disease for participants consuming ≥2 servings/day were 1.21 (95% CI,1.12 to 1.32; *P*-trend<0.001) and 1.03 (95% CI, 0.97 to 1.09; *P*-trend=0.06), respectively. In the joint analyses, for participants meeting and not meeting physical activity guidelines (<7.5 vs ≥7.5 MET-h/week) as well as consuming ≥2 servings/day of SSBs or ASBs, the HRs for cardiovascular disease were 1.15 (95% CI, 1.08 to 1.23) and 0.96 (95% CI, 0.91 to 1.02), and 1.47 (95% CI, 1.37 to 1.57) and 1.29 (95% CI, 1.22 to 1.37) respectively, compared with participants who met physical activity guidelines and never/rarely consumed these beverages. Similar patterns were observed when coronary heart disease and stroke were analyzed.

**Conclusions:** Our findings suggest that among physically active participants, higher SSB intake, but not ASBs, is associated with a higher cardiovascular risk. Our results support current recommendations to limit the intake of SSB and maintain adequate physical activity levels.

**Clinical Perspective:** *What is new?:*

- Consumption of sugar- or artificially-sweetened beverages and physical activity are independently associated with cardiovascular disease; nevertheless, the role that physical activity plays in the associations between sugar- and artificially-sweetened beverages consumption and cardiovascular disease warrants further analysis since physical activity could offset the health risk that would have been induced by their consumption.
- In two US cohorts, not meeting physical activity guidelines jointly with a greater intake of sugar- or artificially-sweetened beverages was associated with a higher risk of cardiovascular disease; however, even when individuals were physically active, higher consumption of sugar-sweetened beverages was associated with a higher cardiovascular risk.

*What are the clinical implications?:*

- Our results provide further evidence of the intake of sugar-sweetened beverages and low physical activity as factors negatively impacting cardiovascular health.
- Our results support public health recommendations and policies to limit the intake of sugar-sweetened beverages and meet and maintain adequate physical activity levels.

## Introduction

Sugar-sweetened beverages (SSBs) have a detrimental impact on health^1–5^ and are associated with a higher risk of cardiovascular disease (CVD),^1,6–11^ the leading cause of mortality worldwide.^12^ Although there has been a nationwide decline in SSB consumption since 2000,^13^ they remain the single largest source of calories and added sugar in the diet.^14^ In contrast, the intake of artificially-sweetened beverages (ASBs) has increased in the United States (US).^15^ They are used as ‘healthier’ alternatives to SSBs and may be an effective strategy to help control caloric intake and support weight loss.^16^ However, evidence on the long-term health impact of ASBs on incident CVD is still inconclusive.^17–19^ Furthermore, the role that physical activity plays in the association between SSB or ASB consumption and the risk of CVD warrants further analysis. Independently, physical activity is cardioprotective,^20,21^ and the majority of SSB or ASB intake studies adjust for physical activity. A counterargument is the probable role that physical activity could play to offset the potential health risk that would have been induced by SSB or ASB intake. Thus, the implication is that given the same level of exposure to SSB or ASB intake, those who meet physical activity recommendations would have a relatively lower risk of CVD than those who are not physically active. Therefore, the aim of this study is to examine the independent and joint associations between SSB or ASB consumption and physical activity in relation to the risk of CVD in men and women from two large US prospective cohort studies.

## Methods

### Study Population and Design

The Nurses’ Health Study (NHS), a prospective study initiated in 1976, enrolled 121,700 registered nurses aged 30-55 years. The Health Professionals Follow-up Study (HPFS), a prospective study established in 1986, enrolled 51,529 US male health professionals aged 40-75 years. Baseline and follow-up questionnaires were sent to participants every 2-years to update medical, lifestyle, and health information. Follow-up rates exceed 90% for each 2-year cycle. Diet was assessed using a self-administered food frequency questionnaire every 4-years, first collected in 1980 in NHS and 1986 for HPFS, used as this study’s baseline cycles. A detailed description of the two cohorts has been previously reported.^22,23^

We excluded individuals with a history of diabetes mellitus, CVD, or cancer at or before baseline; missing information regarding SSB and physical activity; or implausible caloric intake (<500 or >3500 kcal/day in women, and <800 kcal or >4200 kcal/day in men). The final analysis included 65,730 women and 39,418 men. The protocol was approved by the institutional review board of Brigham and Women’s Hospital and Harvard T.H. Chan School of Public Health. All participants gave informed consent.

### Dietary Assessment and Sugar-Sweetened Beverage Intake

Intakes of SSBs and ASBs were assessed using a validated and reproducible food frequency questionnaire designed to assess the usual diet over the previous year. The reproducibility and validity of these instruments have been described previously.^24,25^ Participants were asked how often, on average, they consumed a standard portion of foods and beverages (one standard glass, bottle, or can), with possible responses ranging from ‘never or less than once/month to ‘≥6 times/day’. SSBs were defined as carbonated sugar-sweetened beverages (caffeinated colas, caffeine-free colas, or other), and noncarbonated sugar-sweetened beverages (fruit punches, lemonades, or other fruit drinks). Fruit juices were not considered SSBs since sugar is naturally present. ASBs were defined as caffeinated, caffeine-free, and noncarbonated low-calorie or diet beverages.

### Physical Activity Assessment

Participants’ physical activity was assessed every 4-years by reporting the average time per week spent in various moderate- or vigorous-intensity leisure-time activities in the preceding year, using a validated self-reported questionnaire.^26^ Based on the intensity and duration of each activity, weekly expenditure in metabolic equivalent tasks (MET-hr/week) was calculated and summed over all activities for total physical activity.

### Ascertainment of Cardiovascular Disease Incidence

The primary outcome was incident total CVD, defined as a composite of fatal and nonfatal coronary heart disease (CHD, including myocardial infarction) and fatal and nonfatal stroke. Secondary outcomes were CHD, defined as fatal CHD or nonfatal myocardial infarction, and stroke. In both the NHS and HPFS cohorts, when a participant (or family members) reported an incident event, permission was requested to examine their medical records by the study investigator-physicians who were blinded to the participant’s risk factor status. For each endpoint, the month and year of diagnosis were recorded as the diagnosis date. Nonfatal events were confirmed through a review of medical records. Myocardial infarction was defined according to the World Health Organization criteria.^27^ Strokes were confirmed if data in the medical records fulfilled the National Survey of Stroke criteria.^28^ Strokes were classified as ischemic stroke (thrombotic or embolic occlusion of a cerebral artery), hemorrhagic stroke (subarachnoid and intraparenchymal hemorrhage), or stroke of probable and/or unknown subtype (subtype data not available). Death ascertainment was performed by searching the National Death Index,^29^ by family members’ responses to follow-up questionnaires, or by reports from participants’ professional organizations. We requested access to medical records, autopsy reports, and death certificates to confirm all suspected deaths due to myocardial infarction. Fatal myocardial infarction was confirmed by medical records or autopsy reports. We included all confirmed and probable cases in our report because results were similar after probable cases were excluded. Follow-up for deaths was >98% complete.

### Assessment of Covariates

Information on lifestyle and CVD risk factors was assessed and updated every other year and included: age; weight; smoking status; use of aspirin, multivitamins, postmenopausal hormone-replacement therapy, and oral contraceptives; menopausal status; and hypertension or hypercholesterolemia that had been recently diagnosed by a physician; and family history of chronic diseases. Height was ascertained for women in 1976 and for men in 1986. Height and body weight were used to calculate body mass index (BMI; weight in kilograms divided by height in meters squared). Alcohol intake was assessed, and updated, from the food frequency questionnaires every 4 years. Ancestry information was collected in1986 for men and in 1992 for women. The Alternate Healthy Eating Index (AHEI, range 0-90; higher score indicating adherence to Dietary Guidelines for Americans) was calculated from food frequency questionnaire data, without the exposure (SSBs) and alcohol (we considered as a separate confounder) components, to indicate participant’s overall diet quality.^30^

## Statistical Analysis

Person-years of follow-up were calculated from the return of the baseline questionnaire to the date of diagnosis of CVD, death, or end of follow-up whichever came first. We used Cox proportional-hazards models to estimate hazard ratios and 95% confidence intervals (HRs [CIs]) of developing CVD according to the independent and joint associations of SSB or ASB intakes and physical activity.

For the independent association analysis between SSB or ASB intakes and risk of CVD, we calculated the cumulative averages of SSBs and ASBs, and subtypes, at each updated food-frequency questionnaire to approximate long-term habitual intakes. Beverage intake was categorized by frequency: <1/month (reference), 1–4/month, 2–6/week, 1-<2/day, and ≥2/day, and linear trends were evaluated using the Wald test on a continuous variable representing median intakes of each category. The same categorization was applied to SSB and ASB sub-types. We also evaluated the association for 1 serving increase of SSB and ASB intakes per day in the multivariable models. In Model 1 we adjusted for age; race; ancestry; alcohol intake; smoking status; physical activity (for independent SSB and ASB models, in quintiles); family history of myocardial infarction; baseline hypertension or antihypertensive medication; baseline hypercholesterolemia or cholesterol-lowering medication; multivitamin and aspirin use; menopausal status and use of hormone-replacement therapy (in women); total energy intake (continuous); and BMI (in quintiles). In Model 2 we additionally adjusted for the Alternate Healthy Eating Index (in quintiles, without SSBs and alcohol; not applicable in the independent physical activity models).

We conducted a series of stratified analyses for continuous servings/day of SSBs or ASBs with incidence CVD by pre-specified subgroups and calculated p-values for heterogeneity with Wald tests for the multiplication interaction of SSB or ASB exposures (continuous variable) with the effect modifier variables.

For the joint analyses, SSB or ASB intakes were combined into three categories: (1) never or less than 1 serving/month, (2) 1-4 servings/month, (3) ≥2 servings/week, with physical activity dichotomized according to the current aerobic physical activity guidelines for adults (<7.5 MET-hr/week is equivalent to 150 minutes/week of moderate-intensity physical activity)^31^: <7.5 MET-hr/week vs. ≥7.5 MET-hr/week. We set participants who reported never/rarely consuming SSBs or ASBs and meeting physical activity guidelines as the reference category. We tested the multiplicative interaction with the cross-product interaction term between continuous servings/day of SSB or ASB intakes*total physical activity added to the main effects models. To assess the additive interaction, we derived the probability value of the relative excess risk due to interaction as an index of additive interaction, also with SSB and ASB intakes and physical activity as continuous variables.^32–34^

Because participants may alter dietary patterns after the diagnosis of major illness, we stopped updating dietary variables when participants reported a diagnosis of coronary artery bypass, angina, or cancer, although follow-up continued until CVD endpoint occurrence, death, or the end of the study period.^35^ If food-frequency questionnaire exposure data were missing at a given updated time point, we carried forward the exposure level from the previous cumulative cycles.

### Sensitivity Analyses

We also ran a series of sensitivity analyses to assess the robustness of results in independent and joint associations. First, the diet was continuously updated until the end of follow-up. Second, we excluded BMI from the models. Third, models were adjusted for median household income and education. Fourth, instead of adjusting for AHEI, we adjusted for a set of dietary variables. Lastly, we further examined two sub-groups: 1) ‘heavy SSB or ASB consumers’ (top 25% SSB or ASB consumers vs not) and the dichotomized physical activity variable, and 2) ‘heavy SSB or ASB consumers’ and ‘high physical activity level’ (top 25% physical activity vs not).

All analyses were performed separately for each cohort and then pooled using inverse variance-weighted fixed-effect meta-analysis, and variance-weighted meta-analysis. Statistical tests were two-sided, and *P-*values of < 0.05 were considered to indicate statistical significance. Data were analyzed with the SAS package, version 9.4 (SAS Institute, Cary, NC).

## Results

A total of 13,269 CVD cases (8,438 CHD; 4,997 strokes) were documented, 6,156 in men and 7,113 in women, during a maximum follow-up of 30 (median 13.4) and 36 (median 17.9) years, respectively. Men and women in the reference group, who jointly met physical activity guidelines and consumed less than 1 SSB serving/month at baseline (21% women and 23% men), had on average lower total energy intake and higher diet quality (**Table 1**). In contrast, men and women who did not meet physical activity guidelines and consumed ≥2 servings/week of SSBs at baseline had a higher total energy intake and lower Alternate Healthy Index diet quality score. **Table S1** shows baseline characteristics by categories of SSBs and ASBs intakes separately, and quintiles of physical activity. **Table S2** shows baseline characteristics jointly by categories of ASB and physical activity.

**Table 1.** Baseline^*^ Characteristics of Participants According to Sugar-Sweetened Beverage Intake and Physical Activity Joint Association Categories in Two Large US Cohorts.

|  | Meets the US Physical Activity Guidelines<sup>†</sup> |  |  | Does Not Meet the US Physical Activity Guidelines<sup>†</sup> |  |  |
| --- | --- | --- | --- | --- | --- | --- |
|  | Never or <1 SSB serving per month | 1-4 SSB servings per month | ≥2 SSB servings per week | Never or <1 SSB serving per month | 1-4 SSB servings per month | ≥2 SSB servings per week |
| **Nurses' Health Study** |  |  |  |  |  |  |
| Participants, n | 13,895 | 7,636 | 12,014 | 11,331 | 7,367 | 13,487 |
| Age, years | 47 ± 7 | 47 ± 7 | 45 ± 7 | 48 ± 7 | 47 ± 7 | 45 ± 7 |
| BMI, kg/m<sup>2</sup> | 23.7 ± 3.7 | 23.6 ± 3.8 | 24.0 ± 4.2 | 24.6 ± 4.5 | 24.5 ± 4.4 | 24.9 ± 4.9 |
| Physical activity, MET-h/week | 26.1 ± 24.5 | 24.3 ± 24.2 | 23.9 ± 22.3 | 2.8 ± 2.0 | 2.8 ± 2.0 | 2.8 ± 1.9 |
| Family history of myocardial infarction (%) | 39.6 | 38.5 | 37.5 | 39.5 | 37.7 | 37.1 |
| Family history of diabetes (%) | 28.2 | 27.9 | 27.8 | 30.3 | 29.5 | 30.5 |
| Ethnicity, white (%) | 98.7 | 97.9 | 97.4 | 98.7 | 98.0 | 97.0 |
| Current smoker (%) | 24.3 | 22.1 | 25.7 | 31.9 | 28.3 | 29.8 |
| Hypertension (%) | 5.2 | 4.6 | 5.6 | 6.3 | 5.8 | 6.2 |
| Hypercholesterolemia (%) | 4.1 | 3.6 | 3.6 | 4.2 | 3.6 | 3.6 |
| Multivitamin supplement use (%) | 37.8 | 36.0 | 34.5 | 33.1 | 31.9 | 31.0 |
| Aspirin use (%) | 44.2 | 46.5 | 49.1 | 46.2 | 45.9 | 49.6 |
| Current menopausal hormone use (%) | 7.3 | 6.8 | 6.3 | 7.1 | 6.8 | 6.3 |
| Total energy intake, kcal/day | 1,444 ± 461 | 1,534 ± 470 | 1,718 ± 500 | 1,445 ± 470 | 1,506 ± 473 | 1,705 ± 510 |
| Artificially-sweetened beverages, servings/day | 0.6 ± 1.0 | 0.3 ± 0.7 | 0.3 ± 0.6 | 0.6 ± 1.1 | 0.3 ± 0.7 | 0.3 ± 0.6 |
| Sugar-sweetened beverages, servings/day | 0.0 ± 0.0 | 0.1 ± 0.0 | 0.9 ± 0.9 | 0.0 ± 0.0 | 0.1 ± 0.0 | 1.0 ± 1.0 |
| Total beverages, servings/day | 5.4 ± 2.5 | 5.2 ± 2.4 | 5.7 ± 2.6 | 5.5 ± 2.6 | 5.3 ± 2.5 | 5.8 ± 2.6 |
| Alcohol, g/day | 8.0 ± 11.0 | 6.3 ± 9.6 | 5.5 ± 9.2 | 8.0 ± 12.3 | 5.5 ± 9.9 | 5.0 ± 9.7 |
| Red and processed meat, servings/day | 1.2 ± 0.8 | 1.4 ± 0.8 | 1.5 ± 0.8 | 1.3 ± 0.8 | 1.4 ± 0.8 | 1.6 ± 0.8 |
| Nuts, servings/day | 0.2 ± 0.3 | 0.2 ± 0.3 | 0.2 ± 0.3 | 0.1 ± 0.3 | 0.1 ± 0.3 | 0.1 ± 0.3 |
| Whole grains, servings/day | 2.3 ± 2.0 | 2.2 ± 1.9 | 2.0 ± 1.8 | 2.1 ± 2.0 | 2.1 ± 1.9 | 1.9 ± 1.8 |
| Fruits and vegetables, servings/day | 4.5 ± 2.2 | 4.2 ± 2.0 | 4.1 ± 2.0 | 4.0 ± 2.1 | 3.7 ± 1.9 | 3.6 ± 1.9 |
| AHEI<sup>‡</sup> | 44.6 ± 9.0 | 42.3 ± 8.5 | 40.3 ± 8.3 | 41.2 ± 8.7 | 39.4 ± 8.2 | 37.7 ± 7.8 |
| **Health Professionals Follow-up Study** |  |  |  |  |  |  |
| Participants, n | 9,062 | 5,903 | 10,015 | 4,520 | 3,308 | 6,610 |
| Age, years | 54 ± 9 | 53 ± 9 | 51 ± 9 | 56 ± 9 | 54 ± 9 | 51 ± 9 |
| BMI, kg/m<sup>2</sup> | 25.1 ± 3.1 | 25.2 ± 3.1 | 25.2 ± 3.1 | 26.2 ± 3.7 | 25.9 ± 3.5 | 25.9 ± 3.5 |
| Physical activity, MET-h/week | 32.6 ± 27.3 | 30.4 ± 25.5 | 30.7 ± 26.0 | 3.0 ± 2.2 | 3.1 ± 2.1 | 3.0 ± 2.1 |
| Family history of myocardial infarction (%) | 33.6 | 30.4 | 30.7 | 33.1 | 31.2 | 31.6 |
| Family history of diabetes (%) | 20.0 | 18.4 | 16.9 | 20.3 | 19.2 | 18.3 |
| Ethnicity, white (%) | 95.7 | 96.2 | 94.1 | 96.2 | 95.5 | 94.0 |
| Current smoker (%) | 6.7 | 7.4 | 9.1 | 12.1 | 11.6 | 13.6 |
| Hypertension (%) | 21.2 | 19.2 | 19.6 | 24.5 | 22.9 | 22.5 |
| Hypercholesterolemia (%) | 11.0 | 11.0 | 9.6 | 11.5 | 10.2 | 9.4 |
| Multivitamin supplement use (%) | 46.5 | 43.8 | 41.5 | 41.1 | 37.4 | 36.7 |
| Aspirin use (%) | 25.6 | 25.2 | 27.5 | 27.0 | 26.6 | 27.3 |
| Total energy intake, kcal/day | 1,821 ± 558 | 1,944 ± 575 | 2,219 ± 632 | 1,738 ± 545 | 1,872 ± 556 | 2,163 ± 639 |
| Artificially-sweetened beverages, servings/day | 0.7 ± 1.1 | 0.5 ± 0.8 | 0.3 ± 0.7 | 0.7 ± 1.2 | 0.5 ± 1.0 | 0.3 ± 0.7 |
| Sugar-sweetened beverages, servings/day | 0.0 ± 0.0 | 0.1 ± 0.0 | 0.7 ± 0.7 | 0.0 ± 0.0 | 0.1 ± 0.0 | 0.8 ± 0.8 |
| Total beverages, servings/day | 7.2 ± 3.0 | 7.2 ± 2.8 | 7.7 ± 2.9 | 7.2 ± 3.0 | 7.1 ± 2.9 | 7.4 ± 3.0 |
| Alcohol, g/day | 12.8 ± 16.2 | 11.6 ± 14.7 | 10.8 ± 14.3 | 12.4 ± 16.9 | 11.2 ± 16.0 | 10.0 ± 15.4 |
| Red and processed meat, servings/day | 0.9 ± 0.8 | 1.0 ± 0.8 | 1.3 ± 0.8 | 1.1 ± 0.8 | 1.2 ± 0.8 | 1.4 ± 0.9 |
| Nuts, servings/day | 0.3 ± 0.4 | 0.3 ± 0.4 | 0.3 ± 0.4 | 0.2 ± 0.4 | 0.2 ± 0.4 | 0.2 ± 0.4 |
| Whole grains, servings/day | 1.9 ± 1.6 | 1.8 ± 1.5 | 1.6 ± 1.4 | 1.5 ± 1.4 | 1.5 ± 1.3 | 1.4 ± 1.2 |
| Fruits and vegetables, servings/day | 5.3 ± 2.9 | 5.2 ± 2.9 | 5.1 ± 2.7 | 4.4 ± 2.6 | 4.3 ± 2.4 | 4.3 ± 2.4 |
| AHEI<sup>‡</sup> | 48.2 ± 10.2 | 45.9 ± 9.8 | 43.1 ± 9.4 | 43.9 ± 10.1 | 41.8 ± 9.6 | 39.6 ± 9.1 |

<sup>1</sup>Values are mean ± SD or %, and are standardized to the age distribution of the study population. HPFS, n=39,418; NHS, n=65,730.
<sup>2</sup>\* Baseline for NHS is 1980 and for HPFS is 1986.
<sup>3</sup>† US physical activity guidelines were met if participant reported ≥150 minutes/week of moderate-vigorous physical activity.
<sup>4</sup>‡ AHEI range 0-90, sugar-sweetened beverages and alcohol were removed from the index calculation.
<sup>5</sup>AHEI = alternative healthy eating index; BMI = body mass index; MET = metabolic equivalent task; MVPA = moderate-vigorous physical activity.

**Table 2** shows the pooled associations for intakes of SSBs, ASBs, and physical activity with CVD, CHD, and stroke. In multivariable model 2, compared to participants who never/rarely consumed SSBs, those consuming ≥2 SSB servings/day had a 21% higher risk of CVD (pooled HR 1.21, 95% CI, 1.12 to 1.32; p-trend<0.001). Per serving increment of SSB intake per day, the pooled HR for CVD was 1.15 (95% CI, 1.09 to 1.20; p-trend<0.0001), for CHD 1.17 (95% CI, 1.10 to 1.24; p-trend<0.0001), and for stroke 1.11 (95% CI, 1.02 to 1.20; p-trend=0.02). No evidence of significant differences in ASB intake and risk of CVD or CHD was observed, with the exception of stroke where per serving increment of ASB intake per day was 1.05 (95% CI, 1.01 to 1.10; p-trend=0.03). Comparing the highest to the lowest quintile of physical activity, the pooled hazard ratios were 0.66 (95% CI, 0.63 to 0.70) for CVD, 0.67 (95% CI, 0.63 to 0.72) for CHD, and 0.64 (95% CI, 0.59 to 0.71) for stroke. **Table S3** shows the results for stroke subtypes for SSB, ASB, and physical activity. **Table S4** shows the results for the association between SSB or ASB subtypes and CVD. In the SSB subtype pooled analysis, per serving increment of cola and non-cola carbonated drink intake per day was associated with a15% (pooled HR 1.15, 95% CI, 1.08 to 1.23; p-trend<0.0001) and a 12% (pooled HR 1.12, 95% CI, 1.04 to 1.21; p-trend=0.004) higher risk of CVD, respectively. The associations between SSB, ASB, and physical activity with CVD risks were consistent across subgroups (**Table 3**). Cohort-specific results are shown in **Table S5**.

**Table 2.**
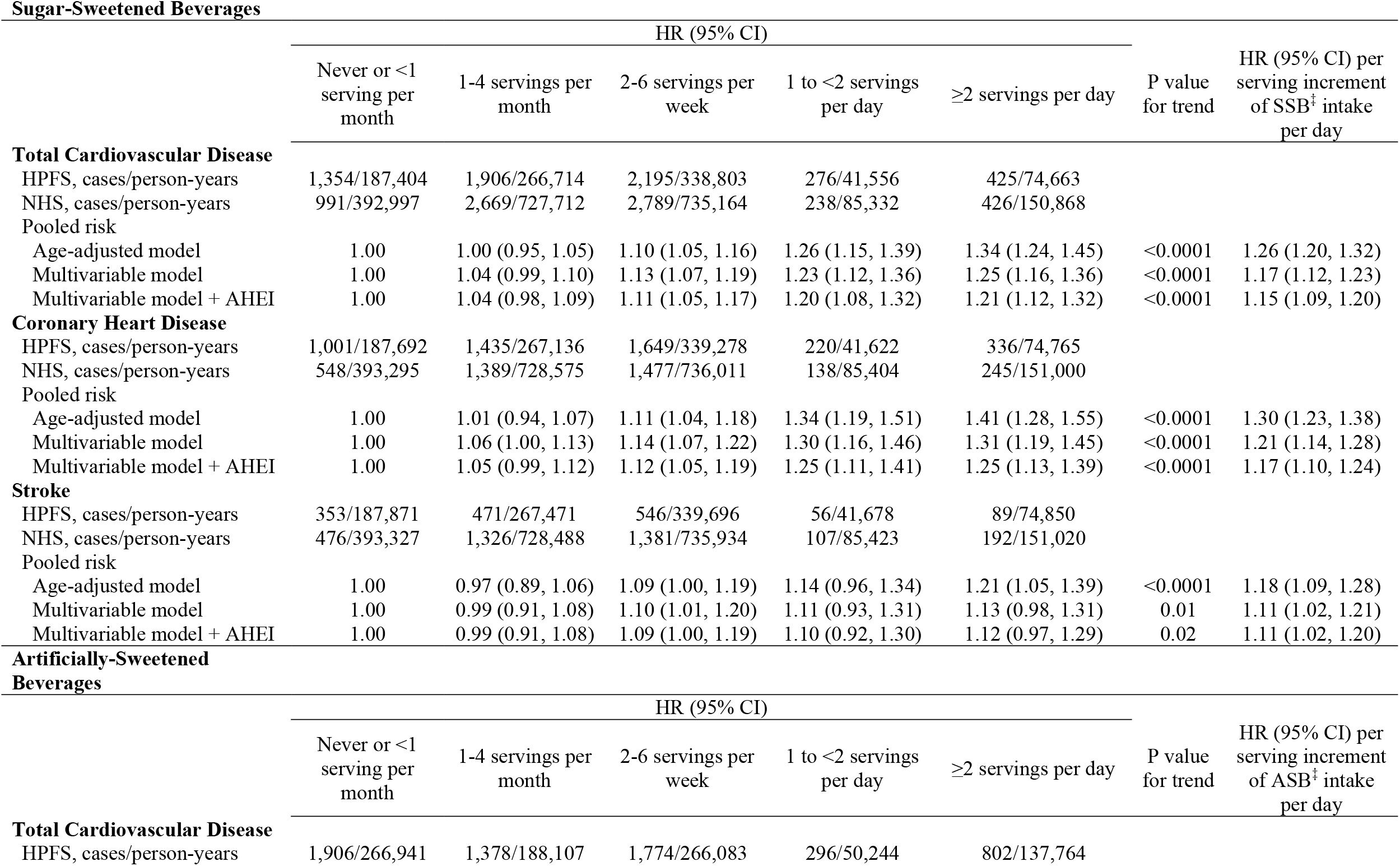
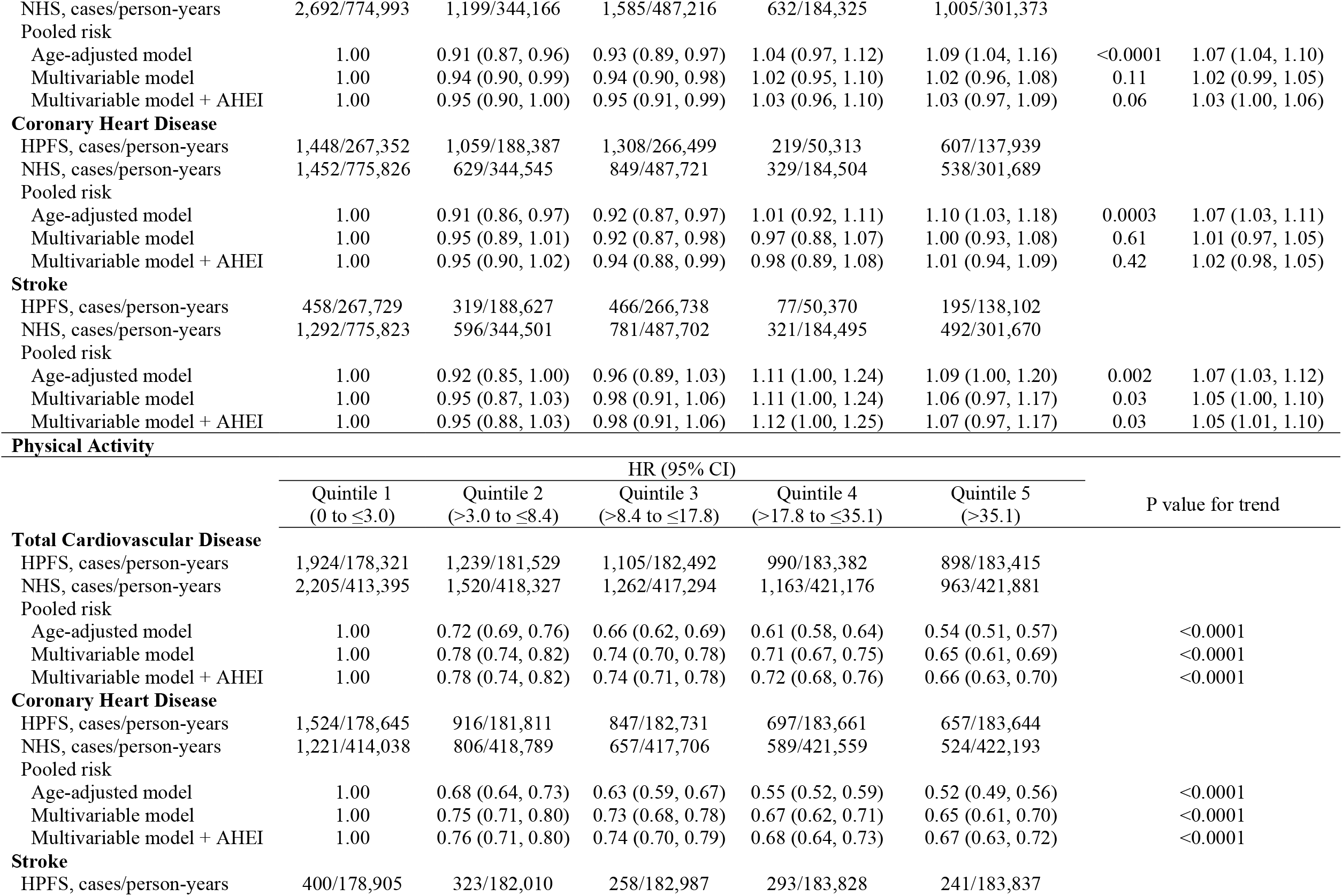
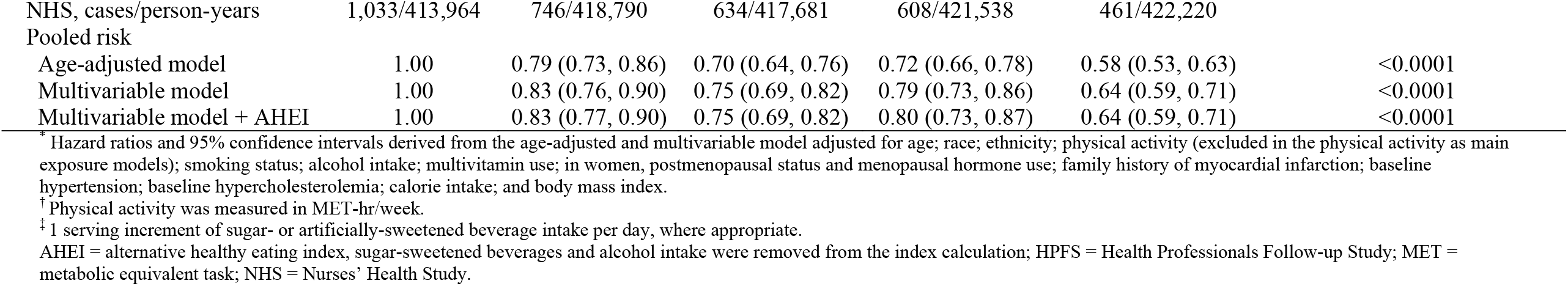
Pooled Risk^*^ of Cardiovascular Events According to Sugar- or Artificially Sweetened Beverage Intake Categories and Physical Activity^†^ Quintiles in Two Large US Cohorts.

**Table 3.** Pooled Subgroup Analyses for Risk^*^ of Cardiovascular Events According to 1 Serving Increment in Sugar- or Artificially-Sweetened Beverage Intake Per Day^†^ in Two Large US Cohorts.

|  | Cardiovascular Disease |  | Coronary Heart Disease |  | Stroke |  |
| --- | --- | --- | --- | --- | --- | --- |
|  | Pooled Adjusted HR (95% CI) | p-value for interaction | Pooled Adjusted HR (95% CI) | p-value for interaction | Pooled Adjusted HR (95% CI) | p-value for interaction |
| **Sugar-Sweetened Beverages** |  |  |  |  |  |  |
| **Age** |  |  |  |  |  |  |
| <65 years | 1.15 (1.06, 1.25) | 0.05 | 1.21 (1.10, 1.34) | 0.04 | 1.01 (0.87, 1.18) | 0.94 |
| ≥65 years | 1.12 (1.05, 1.20) |  | 1.12 (1.03, 1.21) |  | 1.14 (1.03, 1.26) |  |
| **Body mass index, kg/m<sup>2</sup>** |  |  |  |  |  |  |
| <25 | 1.20 (1.12, 1.28) | 0.76 | 1.24 (1.14, 1.35) | 0.45 | 1.12 (0.99, 1.26) | 0.69 |
| ≥25 | 1.09 (1.02, 1.17) |  | 1.09 (1.00, 1.19) |  | 1.10 (0.97, 1.24) |  |
| **Physical activity, meet US guidelines** |  |  |  |  |  |  |
| No (<150 min/week MVPA) | 1.12 (1.04, 1.20) | 0.14 | 1.12 (1.03, 1.23) | 0.07 | 1.09 (0.97, 1.24) | 0.63 |
| Yes (≥150 min/week MVPA) | 1.18 (1.10, 1.26) |  | 1.22 (1.12, 1.32) |  | 1.12 (1.00, 1.26) |  |
| **Television watching, hour/week** |  |  |  |  |  |  |
| <8 | 1.18 (1.08, 1.29) | 0.45 | 1.22 (1.10, 1.36) | 0.59 | 1.12 (0.96, 1.30) | 0.75 |
| 8 to 15.5 | 1.12 (1.00, 1.26) |  | 1.13 (0.98, 1.30) |  | 1.13 (0.93, 1.37) |  |
| ≥15.5 | 1.15 (1.07, 1.24) |  | 1.18 (1.07, 1.29) |  | 1.12 (0.98, 1.26) |  |
| **Smoking status** |  |  |  |  |  |  |
| Never | 1.13 (1.05, 1.21) | 0.01 | 1.16 (1.06, 1.28) | 0.08 | 1.07 (0.94, 1.21) | 0.10 |
| Former/current | 1.20 (1.13, 1.29) |  | 1.21 (1.12, 1.31) |  | 1.19 (1.06, 1.33) |  |
| **Family history of myocardial infarction** |  |  |  |  |  |  |
| No | 1.17 (1.09, 1.25) | 0.89 | 1.19 (1.10, 1.29) | 0.98 | 1.13 (1.02, 1.26) | 0.69 |
| Yes | 1.11 (1.03, 1.20) |  | 1.15 (1.05, 1.26) |  | 1.06 (0.93, 1.22) |  |
| **Calories per day, kcal** |  |  |  |  |  |  |
| <median | 1.19 (1.10, 1.29) | 0.17 | 1.17 (1.06, 1.29) | 0.67 | 1.24 (1.09, 1.41) | 0.06 |
| ≥median | 1.09 (1.02, 1.16) |  | 1.12 (1.03, 1.21) |  | 1.03 (0.92, 1.15) |  |
| **AHEI** |  |  |  |  |  |  |
| <median | 1.13 (1.06, 1.20) | 0.26 | 1.12 (1.04, 1.21) | 0.02 | 1.15 (1.03, 1.28) | 0.34 |
| ≥median | 1.20 (1.11, 1.30) |  | 1.30 (1.18, 1.43) |  | 1.05 (0.92, 1.21) |  |
| **Artificially-Sweetened Beverages** |  |  |  |  |  |  |
| **Age** |  |  |  |  |  |  |
| <65 years | 0.99 (0.93, 1.04) | 0.52 | 0.95 (0.89, 1.01) | 0.16 | 1.08 (0.99, 1.19) | 0.07 |
| ≥65 years | 1.04 (1.01, 1.08) |  | 1.05 (1.00, 1.10) |  | 1.04 (0.99, 1.10) |  |
| **Body mass index, kg/m<sup>2</sup>** |  |  |  |  |  |  |
| <25 | 1.05 (1.01, 1.10) | 0.72 | 1.02 (0.96, 1.08) | 0.46 | 1.10 (1.03, 1.18) | 0.25 |
| ≥25 | 1.03 (0.99, 1.07) |  | 1.04 (0.99, 1.09) |  | 1.02 (0.96, 1.09) |  |
| **Physical activity, meet US guidelines** |  |  |  |  |  |  |
| No (<150 min/week MVPA) | 1.04 (1.00, 1.09) | 0.70 | 1.04 (0.98, 1.10) | 0.60 | 1.05 (0.98, 1.13) | 0.88 |
| Yes (≥150 min/week MVPA) | 1.02 (0.98, 1.06) |  | 1.00 (0.95, 1.05) |  | 1.06 (0.99, 1.13) |  |
| **Television watching, hour/week** |  |  |  |  |  |  |
| <8 | 1.09 (1.04, 1.15) | 0.01 | 1.10 (1.03, 1.18) | 0.004 | 1.08 (1.00, 1.18) | 0.62 |
| 8 to 15.5 | 1.03 (0.96, 1.10) |  | 1.03 (0.95, 1.12) |  | 1.03 (0.93, 1.14) |  |
| ≥15.5 | 0.98 (0.93, 1.03) |  | 0.96 (0.90, 1.02) |  | 1.01 (0.94, 1.09) |  |
| **Smoking status** |  |  |  |  |  |  |
| Never | 1.06 (1.02, 1.11) | 0.0004 | 1.05 (0.99, 1.11) | 0.002 | 1.08 (1.01, 1.16) | 0.10 |
| Former/current | 1.02 (0.98, 1.07) |  | 1.03 (0.97, 1.09) |  | 1.03 (0.96, 1.11) |  |
| **Family history of myocardial infarction** |  |  |  |  |  |  |
| No | 1.03 (0.99, 1.07) | 0.96 | 1.01 (0.96, 1.06) | 1.00 | 1.06 (1.00, 1.13) | 0.98 |
| Yes | 1.03 (0.98, 1.07) |  | 1.02 (0.96, 1.08) |  | 1.04 (0.97, 1.12) |  |
| **Calories per day, kcal** |  |  |  |  |  |  |
| <median | 1.05 (1.01, 1.09) | 0.21 | 1.05 (0.99, 1.10) | 0.10 | 1.06 (1.00, 1.13) | 0.75 |
| ≥median | 1.01 (0.96, 1.05) |  | 0.98 (0.93, 1.04) |  | 1.04 (0.97, 1.11) |  |
| **AHEI** |  |  |  |  |  |  |
| <median | 1.03 (0.98, 1.07) | 0.97 | 1.01 (0.96, 1.07) | 0.99 | 1.05 (0.98, 1.12) | 0.84 |
| ≥median | 1.03 (0.99, 1.07) |  | 1.02 (0.96, 1.07) |  | 1.06 (0.99, 1.13) |  |

\* Hazard ratios and 95% confidence intervals derived from the multivariable model adjusted for age; race; ethnicity; physical activity (excluded in the physical activity as main exposure models); smoking status; alcohol intake; multivitamin use; in women, postmenopausal status and menopausal hormone use; family history of myocardial infarction; baseline hypertension; baseline hypercholesterolemia; calorie intake; body mass index; and AHEI.
† 1 serving of sugar- or artificially-sweetened beverage. HRs for 1 serving increment of sugar- or artificially-sweetened beverage intake per day in each subgroup category.
AHEI = alternative healthy eating index, sugar-sweetened beverages and alcohol intake were removed from the index calculation; MVPA = moderate-vigorous physical activity.

Compared to the reference group (met physical activity guidelines and consumed SSBs <1/month), participants who did not meet the physical activity guidelines and consumed ≥2 servings/week of SSBs, had a pooled HR of 1.47 (95% CI, 1.37 to 1.57) higher risk of CVD (**Figure 1, panel A**). There was also a higher risk of CVD among those who met physical activity guidelines and consumed ≥2 SSB servings/day (pooled HR 1.15, 95% CI, 1.08 to 1.23), with similar patterns for CHD (**Figure 1 panel B**) and stroke (**Figure 1, panel C**). Participants who did not meet physical activity guidelines and consumed ASBs <1/month had a 29% (pooled HR 1.29, 95% CI, 1.22 to 1.37) higher risk of CVD (**Figure 1, panel A**), which was consistent with those who did not meet the physical activity guidelines and consumed ≥2 servings/week of ASBs (pooled HR 1.29, 95% CI, 1.22 to 1.37), compared to the reference group. Cohort-specific results for joint associations are shown in **Table S6**, and stroke subtypes are included in **Table S7. Table S8** shows the results for the joint associations between SSB or ASB subtypes and physical activity and risk of CVD. Although there was no evidence of statistically significant multiplicative interactions between SSB or ASB and physical activity for any outcome, we observed a significant additive interaction between SSB intake and physical activity and risk of CVD (p<0.001) and CHD (p<0.001) (**Table S9**). In addition, we observed a significant additive interaction between the intake of ASBs and physical activity and the risk of CVD (p=0.03) and stroke (p=0.04).

**Figure 1.**
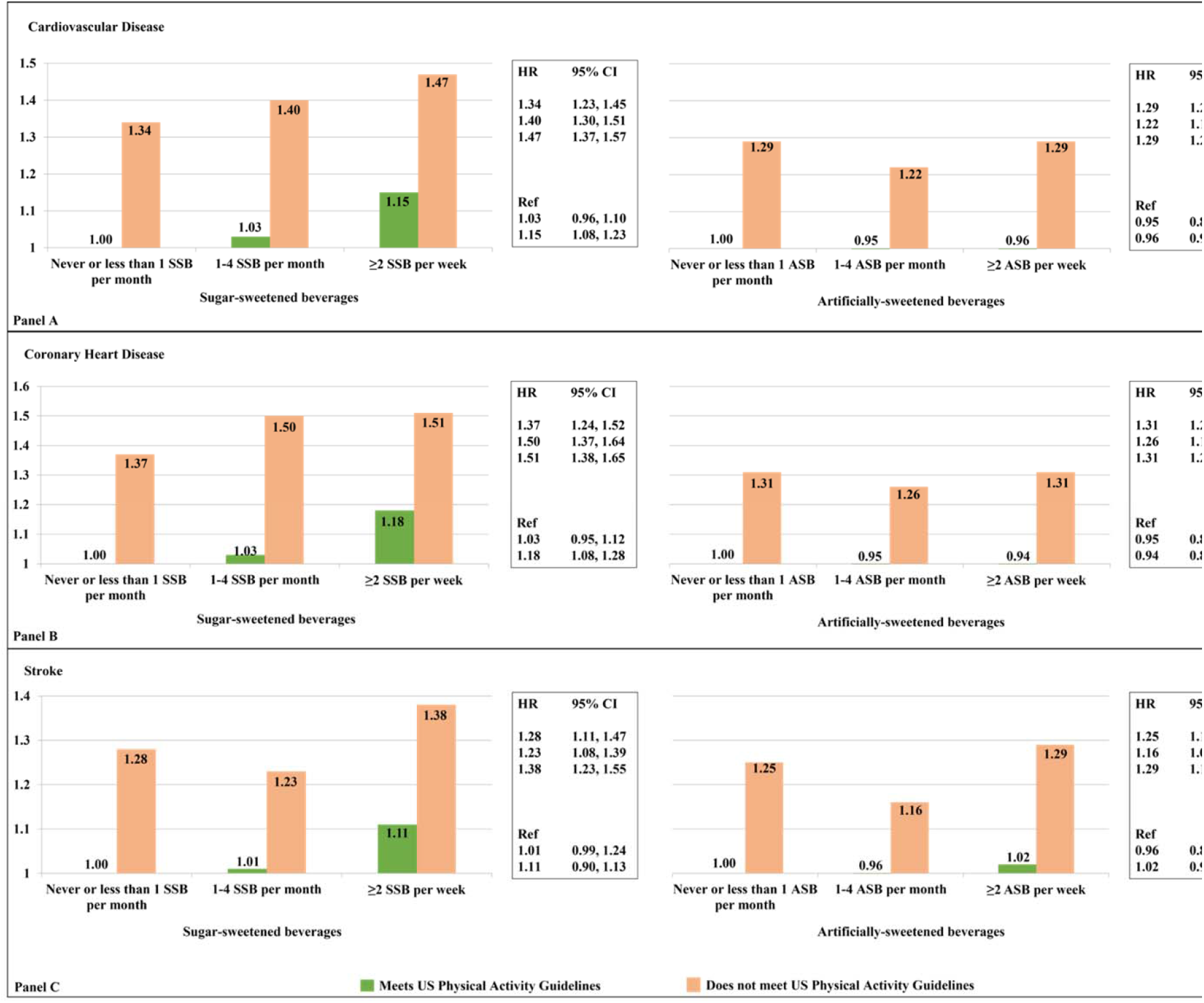
Pooled Risk ^*^ of Cardiovascular Events Associated with the Joint Associations of Sugar- or Artificially-Sweetened Beverages and Physical Activity^†^ in Two Large US Cohorts. ^*^ Hazard ratios and 95% confidence intervals derived from the multivariable model adjusted for the following: age, ethnicity/race; smoking status; alcohol intake; multivitamin use; in women, postmenopausal status and menopausal hormone use; family history of myocardial infarction; baseline hypertension; baseline hypercholesterolemia; calorie intake; body mass index; and AHEI. ^†^ Physical activity was dichotomized to meet or do not meet the US Physical activity guidelines of ≥150 minutes/week of moderate-vigorous physical activity recommendations (corresponds to ≥7.5 MET hr/week or ≥450 MET min/week). AHEI = alternative healthy eating index, sugar-sweetened beverages and alcohol intake were removed from the index calculation; MET = metabolic equivalent task.

The results of these analyses were consistent with those from primary analyses. When we selectively stop updating diet, the independent associations between SSB or ASB consumption and risk of CVD and CHD, were slightly attenuated, while the independent association between SSB intake and risk of stroke remained consistent (**Table S10**). The results of the independent association between physical activity and risk of CVD remained unchanged (**Table S10**). The SSB or ASB intakes and physical activity joint associations and risk of CVD findings were consistent after we stopped updating diet (**Table S11**). The independent and joint association results persisted after we excluded BMI from the final model (**Tables S12 and S13**), except for the ASBs and physical activity joint association results that were strengthened (**Table S13**). Results remained consistent when we additionally adjusted for socioeconomic status (**Tables S14 and S15**) and when we adjusted for dietary factors in lieu of AHEI (**Tables S16 and S17**). The pooled hazard ratios for risk of CVD in ‘heavy SSB consumers’ who met and did not meet the physical activity guidelines were 1.14 (95% CI, 1.08 to 1.21) and 1.44 (95% CI, 1.35 to 1.53), respectively (**Table S18**). The pooled hazard ratios for those ‘heavy SSB or ASB consumers’ and that were in the top 25% of physical activity (‘high physical activity level’) were slightly attenuated - vs using dichotomized physical activity - for all outcomes (**Table S19**).

## Discussion

In two large prospective cohorts of US men and women followed for more than 30 years, we found positive linear associations between SSB consumption and the risk of CVD and CHD, while physical activity was inversely associated with CVD, CHD, and stroke risk, after adjusting for risk factors and other dietary variables. Marginally significant associations were observed per serving increment/day of ASB intake and the risk of CVD. Most importantly, our joint analysis showed that individuals who met physical activity guidelines and consumed >2 servings/day of SSBs had a higher risk of CVD, compared to those who met physical activity guidelines and consumed SSBs <1/month. To our knowledge, the present study is the first large prospective study to examine the joint associations of SSB or ASB consumption and physical activity and the incidence of CVD. Overall, our study suggests that the protective effects of physical activity do not mitigate the detrimental impact of SSB consumption on cardiovascular health.

Findings from several meta-analyses have consistently shown that SSB intake is associated with CVD risk in adults, after adjustment for physical activity.^3,5,36^ The most recent meta-analysis (seven prospective cohorts with 16,937,316 person-years of follow-up and 16,915 incident CVD cases), found that SSB consumption was associated with a 9% higher risk of CVD (95% CI, 1.01 to 1.18) when extreme categories of intake (none or less than one/month vs one or more/day) were compared.^5^

In regard to ASBs, a meta-analysis including six prospective studies (16,281,005 person-years of follow-up with 18,077 incident CVD events) showed that per serving increment of ASBs per day was associated with a 7% higher risk of incidence of CVD (95% CI, 1.05 to 1.10).^5^ Additionally, per serving increase of ASBs per day was associated with a 6% higher risk of CHD and a 9% higher risk of stroke. The dose-response results of our study were weaker because we observed a 3% higher risk of CVD, with marginal significance, and a 5% higher risk of stroke.

Concerning physical activity, we observed a 34% lower risk in the incidence of CVD in those in the highest quintile of physical activity, when compared to the lowest quintile and after accounting for demographic and lifestyle factors. A consistent and strong protective association has been reported with leisure time (relative risk 0.76, 95% CI, 0.70 to 0.82 in men 0.73, 95% CI, 0.68 to 0.78 in women) and occupational physical activity (relative risk 0.89, 95% CI, 0.82 to 0.97 in men and 0.83, 95% CI, 0.67 to 1.03 in women).^37^ Furthermore, a recent systematic review and meta-analysis (33 studies with 1,683,693 participants and 89,493 events, follow-up=12.8 years) reported that an increase from being inactive to attaining recommended physical activity levels was associated with a lower risk of CVD incidence by 17% (95% CI, 0.77 to 0.89), after adjusting for body weight.^38^

The joint associations between SSB or ASB intakes and physical activity and risk of CVD findings are novel. Findings provide evidence against the argument that physical activity could counterbalance the potential health risk that would have been induced by SSB or ASB intake. Even though participants who did not meet the physical activity recommendations had a higher risk of CVD at all levels of SSB or ASB intakes, consuming ≥2 servings SSBs/week, compared to never/rarely consumers of SSBs, while meeting physical activity guidelines was associated with a 15% higher risk of CVD. Although we did not observe a multiplicative interaction between SSB or ASB and physical activity and risk of CVD, we did detect significant additive interactions between SSB and physical activity and risk of CVD and CHD; and ASB and physical activity and risk of CVD and stroke. Of note, in a previous study in men, De Koning et al., reported no significant interaction between SSB and physical activity.^8^ Hence, meeting physical activity recommendations does not modify the CVD risk associated with higher SSB intake. Interestingly, the impact of SSBs on CVD risk is still observed even in participants who met physical activity recommendations and were in the top 25% of physical activity levels.

Strengths of this study include large sample size, prospective population-based design with long and high rates of follow-up, well-defined outcomes, and the use of repeated and validated measurements of diet and other lifestyle data. Limitations of this study include the difficulty in inferring causality due to the observational design. Additionally, residual confounding remains a possibility even though the analyses were extensively adjusted for potential confounders. As well, reverse causation in the ASB analyses is a plausible limitation. Moreover, the dietary information was self-reported and our assessment of SSBs and ASBs and physical activity inevitably had some degree of measurement error. However, the use of repeated measurements lessens random measurement error caused by within-person variation. Also, our study population consisted of primarily non-Hispanic white health professionals, thus the generalizability of our results to other populations may be limited.

## Conclusion

Consumption of SSBs or ASBs and physical activity are independently associated with the risk of CVD. Not meeting physical activity guidelines jointly with a greater intake of SSBs or ASBs was associated with a higher risk of CVD. Additionally, findings from this study suggest that even when individuals are physically active, higher consumption of SSBs is associated with a higher risk of CVD. Our results underscore the adverse health effects of both SSBs consumption and being physically inactive and support public health recommendations and policies to limit the intake of SSBs and maintain adequate physical activity levels.

## Data Availability

The data that support the findings of this study are available from the corresponding author upon reasonable request.

## Acknowledgements

The authors would like to thank the participants and staff of the NHS and HPFS cohorts for their continuous involvement and valuable contributions.

